# DNA methylation aging clocks and pancreatic cancer risk: Pooled analysis of three prospective nested case-control studies

**DOI:** 10.1101/2020.01.30.20019174

**Authors:** Mei Chung, Mengyuan Ruan, Naisi Zhao, Devin C. Koestler, Immaculata De Vivo, Karl T. Kelsey, Dominique S. Michaud

## Abstract

**Background:** Pancreatic cancer is projected to become the second leading cause of cancer-related death by 2030 in the United States. DNA methylation (DNAm) age may reflect age-related variations in the biological changes and abnormalities related to cancer development.

**Method:** We conducted a pooled analysis using prediagnostic blood samples of pancreatic cancer cases and matched controls selected from the Nurses’ Health Study (NHS), the Physician’s Health Study (PHS), and the Health Professionals Follow-up Study (HPFS). We used three DNAm aging clocks (Hannum, Horvath, and PhenoAge) to estimate subjects’ DNAm age, epigenetic age acceleration (AA) and intrinsic epigenetic age acceleration (IEAA) metrics. We performed conditional logistic regression and multivariable Cox proportional hazard regression to examine associations between six AA and IEAA metrics and risk of pancreatic cancer and survival, respectively.

**Results:** A total of 393 incidence pancreatic cancer cases and 431 matched controls from the NHS, PHS, and HPFS cohorts were included in this analysis. The medians of all three epigenetic AA and three IEAA metrics were consistently above zero (indicating accelerated age) among cases, while they were below zero (indicating decelerated age) among the matched controls. Comparing participants in the highest quartile of age acceleration metrics, the pancreatic cancer risks were significantly increased by 67% to 83% for Hannum and PhenoAge AA or IEAA metrics with minimal of 7- to 9-years accelerated ages. Except for Hovarth AA and IEAA metrics, there were significant dose-response trends, such that higher age accelerations were associated with higher pancreatic cancer risk, but the relationships were nonlinear. Stratified analyses showed heterogeneous associations, varying by participants’ characteristics and by epigenetic AA or IEAA metrics. As time to diagnosis increased, the ORs of pancreatic cancer for the Hannum AA and Horvath AA or IEAA metrics trended upwards, while the ORs for the PhenoAge AA or IEAA and Hannum IEAA metrics trended downward. Overall, we observed no significant association between pancreatic cancer survival and any of the prediagnostic epigenetic AA or IEAA metrics.

**Conclusion:** Our results indicate DNAm age acceleration is associated with an increased risk of pancreatic cancer in a nonlinear, dose-response manner. Epigenetic IEAA metrics may be a useful addition to current methods for pancreatic cancer risk prediction.

## Background

DNA methylation (DNAm) clocks are derived from epigenetic DNAm markers that are strongly correlated with chronological age or time and can accurately quantify an age-related phenotype or outcome, or both (1). DNA methylation aging clocks capture age-related epigenetic variations, which can be divided into intrinsic (or intra-cellular) and extrinsic (or broadly within-tissue and external) aspects of the aging process (2). Intrinsic DNAm age is independent of age-related changes in blood-cell composition, while extrinsic DNAm age incorporates age-related changes in blood cell composition in the clocks’ algorithms.

Deviations between DNAm age and chronological age are commonly referred to as “age acceleration,” or positive deviation between DNAm age and chronological age. Numerous cohort studies have reported that subjects with age acceleration were at an increased risk for all-cause mortality after controlling for known risk factors (2-7). Furthermore, a study showed that the offspring of semi-supercentenarians (subjects who reached an age of 105-109 years) have a lower epigenetic age than age-matched controls (8). A meta-analysis of six longitudinal cohorts found that DNAm age increases at a slower rate than chronological age across the life course, and the likely explanation for this phenomenon is survival bias, where healthy individuals are those maintained within a longitudinal study (9). Based on these findings, it has been hypothesized that epigenetic processes play a role in healthy aging, and DNAm age captures some aspects of biological age. DNAm is thus thought to be a marker of susceptibility to disease and is associated with multiple health outcomes (10).

Pancreatic cancer incidence rates have been steadily increasing since 2010, and pancreatic cancer is projected to become the second leading cause of cancer-related death by 2030 in the United States (11). DNAm age may reflect age-related variations in the biological changes and abnormalities related to cancer development. Observational studies have reported that age acceleration estimated using different DNAm clocks is associated with increased all cancer risk and shorter cancer survival independent of major health risk factors (12, 13).

Consistent findings have been reported in studies that examined the association between DNAm age acceleration and specific type of cancer. In a pooled analysis of seven cohort studies, age acceleration had stronger associations with the risk of kidney cancer and B-cell lymphoma than colorectal, gastric, lung, prostate, and urothelial cancer (12). Intrinsic DNAm age acceleration estimated by Horvath’s clock (Horvath IEAA) was significantly associated with an increased risk of lung cancer in the Women’s Health Initiative (WHI) cohort (14). Horvath IEAA was also associated with an increased risk of breast cancer in a nested case-control study embedded in the European Prospective Investigation into Cancer and Nutrition (EPIC) cohort (15). Consistently, in a recent case-cohort study, significant positive associations were observed between age acceleration estimated by three different DNAm clocks (Hannum, Horvath, and PhenoAge) and risk of breast cancer (16).

To our knowledge, no study has examined epigenetic clocks in relation to pancreatic cancer risk or survival. Here, we assessed the associations of DNAm age acceleration and pancreatic cancer risk using incident pancreatic cancer cases and matched controls identified from three large cohort studies in the United States.

## Methods

### Study sample and blood collection

A pooled analysis using prediagnostic bloods of pancreatic cancer cases and matched controls selected from the Nurses’ Health Study (NHS) (17), the Physician’s Health Study (PHS) (18), and the Health Professionals Follow-up Study (HPFS) (19) was conducted. All blood specimens (for each cohort) were drawn by participants and mailed overnight, and upon receipt, the samples were aliquotted and frozen for future analyses.

### Pancreatic cancer cases and matched controls

For any report of cancer (except basal cell skin cancer) in the three cohorts, written permission from study participants or next-of-kin was obtained to review their medical records. In the case permission could not be obtained, cancer registries were used to obtain details of diagnosis. Non-respondents are telephoned to obtain verbal confirmation of the information reported on the follow-up questionnaire (probing for details of diagnosis and treatment), and the social security death index is used to monitor death for remaining non-respondents. The active surveillance for mortality ensures virtually complete ascertainment of cancer deaths. All medical records were reviewed by trained physicians. The report is classified as confirmed cancer only if confirmed by the pathology report.Over 90% of participants have granted permission for medical record review.

During follow-up (through 2018 for HPFS and NHS, and through 2010 for PHS), 403 incident cases were confirmed to have pancreatic cancer among the participants who provided blood samples. A control subject was matched to each case on cohort (which also matches on sex), age (+/- 1 year), date of blood draw (month 3+/- and year), smoking (never, former, current) and race (White/other). Due to low DNA concentrations in some of the samples, and samples removed after data processing of the arrays (see Supplemental Methods), the initial 1:1 matching was not always conserved and resulted in some cases and controls with no matched pair. The final dataset consisted of 393 incidence pancreatic cases and 431 matched controls from the three large cohort studies where blood samples had been collected between 6 months and 26 years prior to cancer diagnosis.

### DNA extraction and bisulfite conversion, and DNA methylation processing

DNA extracted from buffy coats were bisulfite-treated and DNA methylation was measured with the Illumina Infinium MethylationEPIC BeadChip [850K] arrays (Illumina, Inc, CA, USA). Details on DNA methylation measurements and data processing are provided in the Supplementary Methods.

### Methylation age and age acceleration estimation

Three DNAm clocks (Hannum, Horvath, and PhenoAge) were used to estimate subjects’ DNAm age and age acceleration. All three DNAm clocks were developed based on Illumina 450K arrays; some CpGs from the 450K arrays were missing in the most updated Illumina EPIC [850K] arrays but the DNAm age estimates are highly correlated between the two arrays (*r* > 0.91) (20). The missing CpG values were imputed using the ENmix Bioconductor package (21). The Hannum’s clock is comprised of 71 CpG that strongly capture changes in chronological age, which is partly driven by age-related shifts in blood cell composition (22). The DNAm clock algorithm developed by Horvath was constructed across multiple tissues and included 353 CpGs (23). The more recent PhenoAge clock, developed by Levine and colleagues, was trained on age-related and disease phenotypes in combination with chronological age. It incorporates age-related biochemical measures and included 513 CpGs (24). Of the 513 CpGs, 41 and 6 CpGs were overlapped with Horvath’s clock and Hannum’s clock, respectively.

For each of the three DNAm clocks, DNAm age acceleration (AA) was defined by regressing DNAm age on chronologic age and calculating the difference between the observed chronological age and the fitted DNAm age (i.e., the residual). Additionally, intrinsic epigenetic age acceleration (IEAA) metrics were calculated using the residuals from the linear regression of DNAm age on the chronologic age and the measures of blood cell compositions. For all age acceleration metrics, a positive value indicates that DNAm age is higher than expected given the individual’s chronological age (accelerated aging), whereas a negative value indicates that DNAm age is lower than expected given the individual’s chronological age (decelerated aging). Subjects with the absolute value of the age acceleration estimate greater than three standard deviations (SDs) were excluded.

### Statistical analysis

All statistical analysis was performed in R (version 3.6.1). Spearman’s rank correlation was used to calculate the correlations between age acceleration metrics. Results were plotted using *ggplot2*.

To examine association with pancreatic cancer, conditional logistic regression was performed, given that our samples were matched by cohorts (HPFS, NHS, or PHS), age, date of blood draw, race and smoking status using an incident sampling design. Body mass index (BMI) was adjusted in all conditional logistic regression models. Quartiles were assigned according to distribution of each DNAm age acceleration among controls. Similar results were observed when using unconditional logistic regression models including unmatched cases and controls (Supplemental Table 1).

Stratified analyses were also performed to explore whether odd ratios differed by sex, age groups (≤ 65 or >65 years), time to cancer diagnosis (≤10, 10-15, or >15 years), smoking status (current, former, or never smokers), and BMI categories (<25, ≥25 kg/m^2^). To reduce the number of group comparisons, participants were grouped into two groups, i.e., epigenetically older (values of AA or IEAA metrics ≤ 0) versus epigenetically younger (values of AA or IEAA metrics > 0), in these analyses. All stratified analysis models adjusted for BMI, except for the analysis by BMI categories.

The association between survival time and age acceleration among pancreatic cancer cases was examined using multivariable Cox proportional hazard model. Covariates included in the model were cohorts, age at blood draw, race, smoking status, and BMI. Subjects with survival time equal to zero were excluded because these subjects had no medical records and were identified using the National Death Index. Participants with missing covariate data were excluded from the analyses.

## Results

### Study population

Participants in the nested case-control study were diagnosed with pancreatic cancer an average of 13 years (range 6 months to 26 years) after providing a blood sample. About 45% of cases/controls were women (mean age = 59.4), all selected from the NHS cohort. The remaining cases/controls were men from the PHS (mean age = 56.5) and HPFS (mean age = 63.7) cohorts. The descriptive statistics of Hannum, Horvath, and PhenoAge epigenetic metrics by the characteristics of the study participants are provided in Table 1. About 40% of the study participants were never smokers, 45% former smokers, and 15% current smokers. About half of the study participant were overweight or obese. For all three AA metrics, there were more pancreatic cancer cases with accelerated epigenetic age than the matched controls. There were fewer female participants had accelerated epigenetic age than male.

**Table 1.** Baseline characteristics for study population, stratified by epigenetic age acceleration or deceleration^a^

| Mean (SD) or *n* (%) | Overall | Hannum AA |  | Horvath AA |  | PhenoAge AA |  |
| --- | --- | --- | --- | --- | --- | --- | --- |
|  |  | Deceleration | Acceleration | Deceleration | Acceleration | Deceleration | Acceleration |
| *n* | 824 | 407 | 407 | 405 | 411 | 399 | 419 |
| **Pancreatic cancer cases<sup>b</sup>** | 393 (47.7%) | 182 (44.7%) | 203 (49.9%) | 184 (45.4%) | 204 (49.6%) | 178 (44.6%) | 211 (50.4%) |
| **Cohort** |  |  |  |  |  |  |  |
| NHS | 370 (44.9%) | 210 (51.6%) | 160 (39.3%) | 203 (50.1%) | 166 (40.4%) | 186 (46.6%) | 184 (43.9%) |
| HPFS | 297 (36.0%) | 128 (31.4%) | 159 (39.1%) | 134 (33.1%) | 157 (38.2%) | 137 (34.3%) | 154 (36.8%) |
| PHS | 157 (19.1%) | 69 (17.0%) | 88 (21.6%) | 68 (16.8%) | 88 (21.4%) | 76 (19.0%) | 81 (19.3%) |
| **Age at blood draw, year** | 60.4 (7.8) | 59.3 (7.9) | 61.3 (7.5) | 59.5 (8.0) | 61.1 (7.5) | 59.7 (8.0) | 61.0 (7.6) |
| **Time before diagnosis, year<sup>c</sup>** | 13.0 (6.2) | 13.1 (6.4) | 12.9 (6.1) | 13.1 (6.5) | 12.9 (6.0) | 13.1 (6.3) | 12.8 (6.2) |
| **Female** | 370 (44.9%) | 210 (51.6%) | 160 (39.3%) | 203 (50.1%) | 166 (40.4%) | 186 (46.6%) | 184 (43.9%) |
| **White<sup>d</sup>** | 752 (94.5%) | 365 (92.9%) | 377 (95.9%) | 365 (92.6%) | 380 (96.2%) | 367 (94.8%) | 379 (94.0%) |
| **Smoking status<sup>e</sup>** |  |  |  |  |  |  |  |
| Never | 332 (40.6%) | 159 (39.3%) | 169 (41.8%) | 158 (39.4%) | 171 (41.7%) | 166 (42.0%) | 164 (39.3%) |
| Former | 364 (44.5%) | 181 (44.7%) | 178 (44.1%) | 179 (44.6%) | 181 (44.1%) | 171 (43.3%) | 189 (45.3%) |
| Current | 122 (14.9%) | 65 (16.0%) | 57 (14.1%) | 64 (16.0%) | 58 (14.1%) | 58 (14.7%) | 64 (15.3%) |
| **BMI<sup>f</sup>, kg/m<sup>2</sup>** | 25.8 (4.1) | 26.0 (4.2) | 25.7 (4.0) | 25.7 (4.1) | 26.0 (4.1) | 25.6 (3.9) | 26.1 (4.3) |
| Normal and lower (BMI (<25) | 398 (49.1%) | 197 (48.9%) | 197 (49.6%) | 205 (51.5%) | 191 (47.3%) | 205 (51.9%) | 190 (46.5%) |
| Overweight (BMI 25 to <30) | 295 (36.4%) | 146 (36.2%) | 143 (36.0%) | 139 (34.9%) | 150 (37.1%) | 143 (36.2%) | 149 (36.4%) |
| Obese (BMI ≥30) | 117 (14.4%) | 60 (14.9%) | 57 (14.4%) | 54 (13.6%) | 63 (15.6%) | 47 (11.9%) | 70 (17.1%) |
| **Diabetes<sup>g</sup>** | 30 (3.6%) | 13 (3.2%) | 17 (4.2%) | 15 (3.7%) | 15 (3.6%) | 12 (3.0%) | 18 (4.3%) |

AA = epigenetic age acceleration metric; BMI = body mass index; HPFS = Health; Professionals Follow-up Study; IEAA = intrinsic epigenetic age acceleration metric; *n* = sample size; NHS = Nurses' Health Study; PHS = Physicians' Health Study; SD = standard deviation.
<sup>a</sup> Subjects with the absolute value of the age acceleration estimate greater than three SDs were excluded.
<sup>b</sup> Case status based the end of the cohort (other covariates based on questionnaires closest to time of blood draw).
<sup>c</sup> 11 missing values; <sup>d</sup> 28 missing values; <sup>e</sup> 6 missing values; <sup>f</sup> 14 missing values; <sup>g</sup> 1 missing values.

The correlations between the three DNAm age and chronological age were moderate for all three epigenetic clocks (*r* = 0.42, 0.40, and 0.32 for Hannum, Horvath, and PhenoAge, respectively) (Figure 1). Correlations between each pair of age acceleration (AA) or intrinsic age acceleration (IEAA) metrics were high (*r* ranged from 0.65 to 0.98) in our study sample (Figure 2). The distributions of epigenetic AA and IEAA metrics among pancreatic cancer cases and their matched controls are shown in Figure 3. Consistently, the medians of all three epigenetic AA and three IEAA metrics were above zero (indicating accelerated age) among pancreatic cancer cases, while they were below zero (indicating decelerated age) among the matched controls.

**Figure 1:**
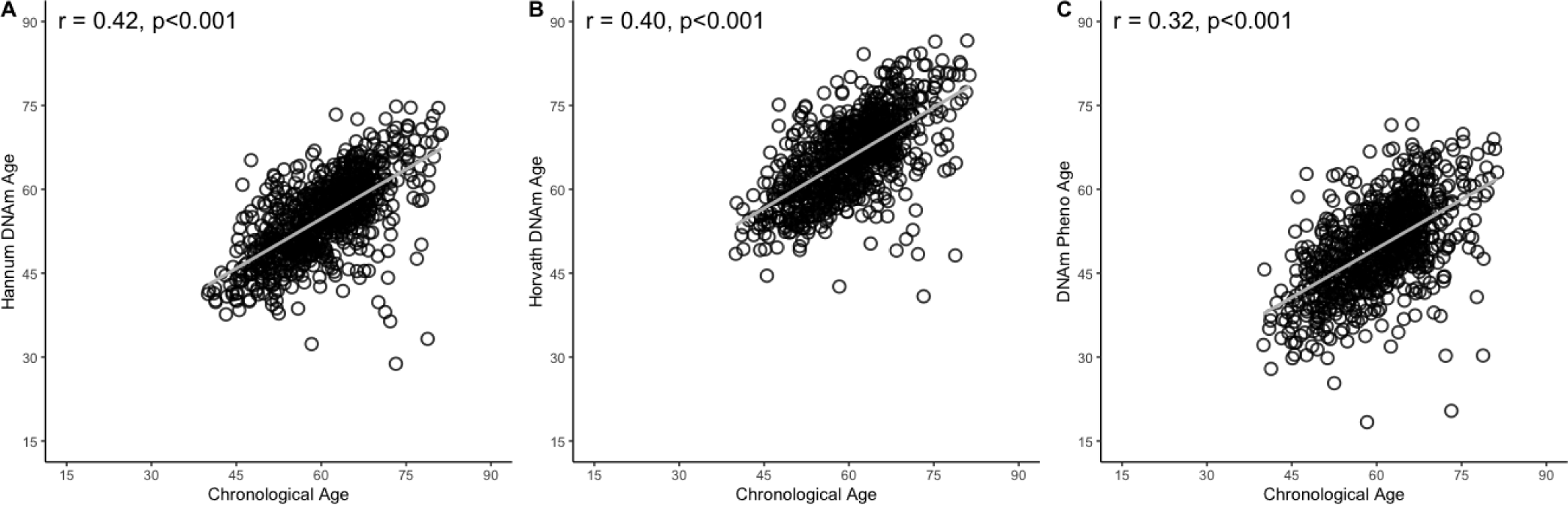
Correlations between chronological age and DNAm age estimated by three epigenetic clocks. Legends: Scatterplots with Pearson correlation coefficients (*r*) and p-values for the relationship between chronological age and DNA methylation age estimated by the Hannum (panel A), Horvath (panel B) and PhenoAge (panel C) clocks.

**Figure 2:**
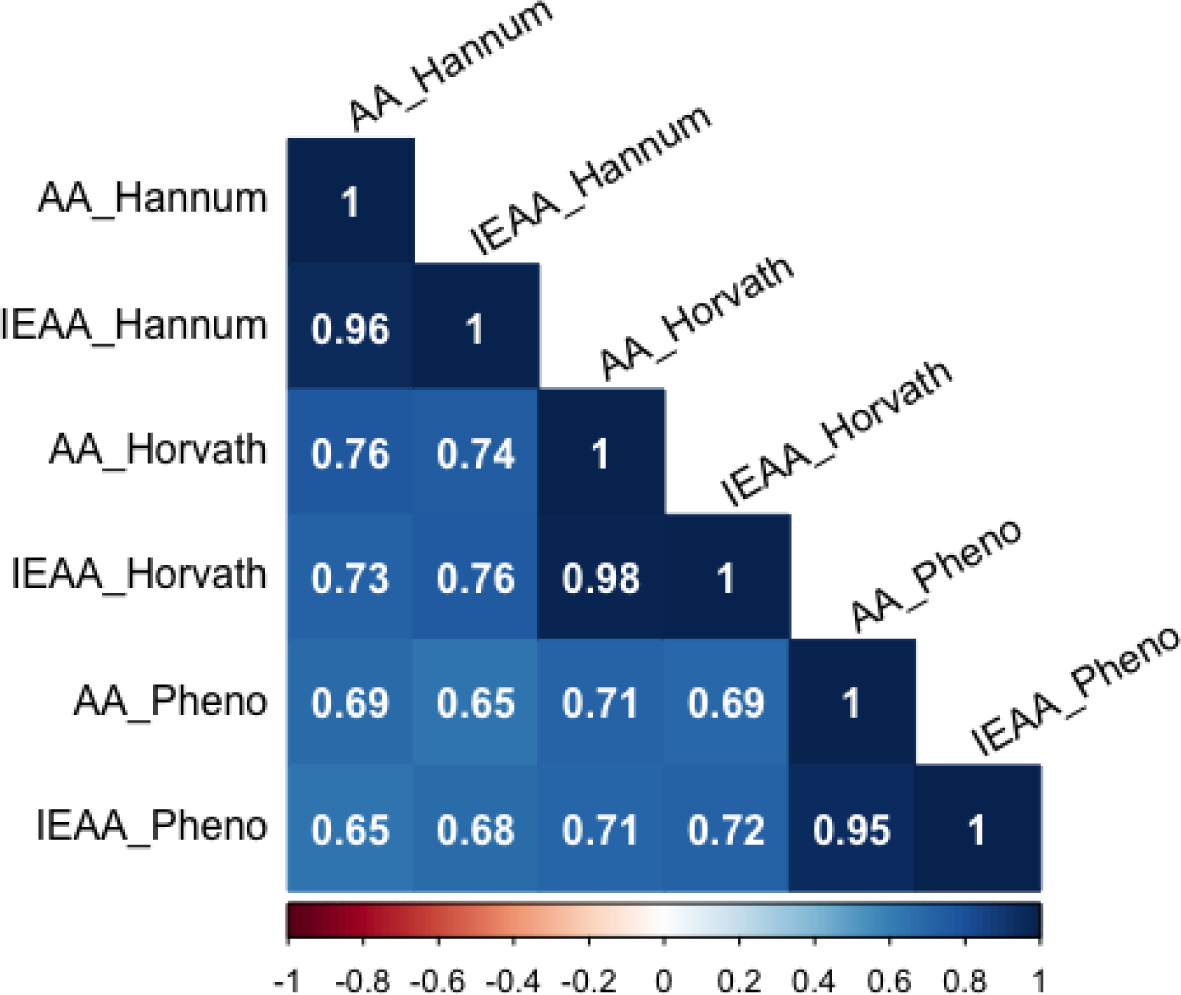
Correlation matrix of age acceleration metrics estimated by three epigenetic clocks. Legends: Spearman’s rank correlations between each pair of epigenetic age acceleration (AA) or intrinsic epigenetic age acceleration (IEAA) metrics estimated by Hannum clock, Horvath clock, and PhenoAge are shown in the boxes of the matrix.

**Figure 3:**
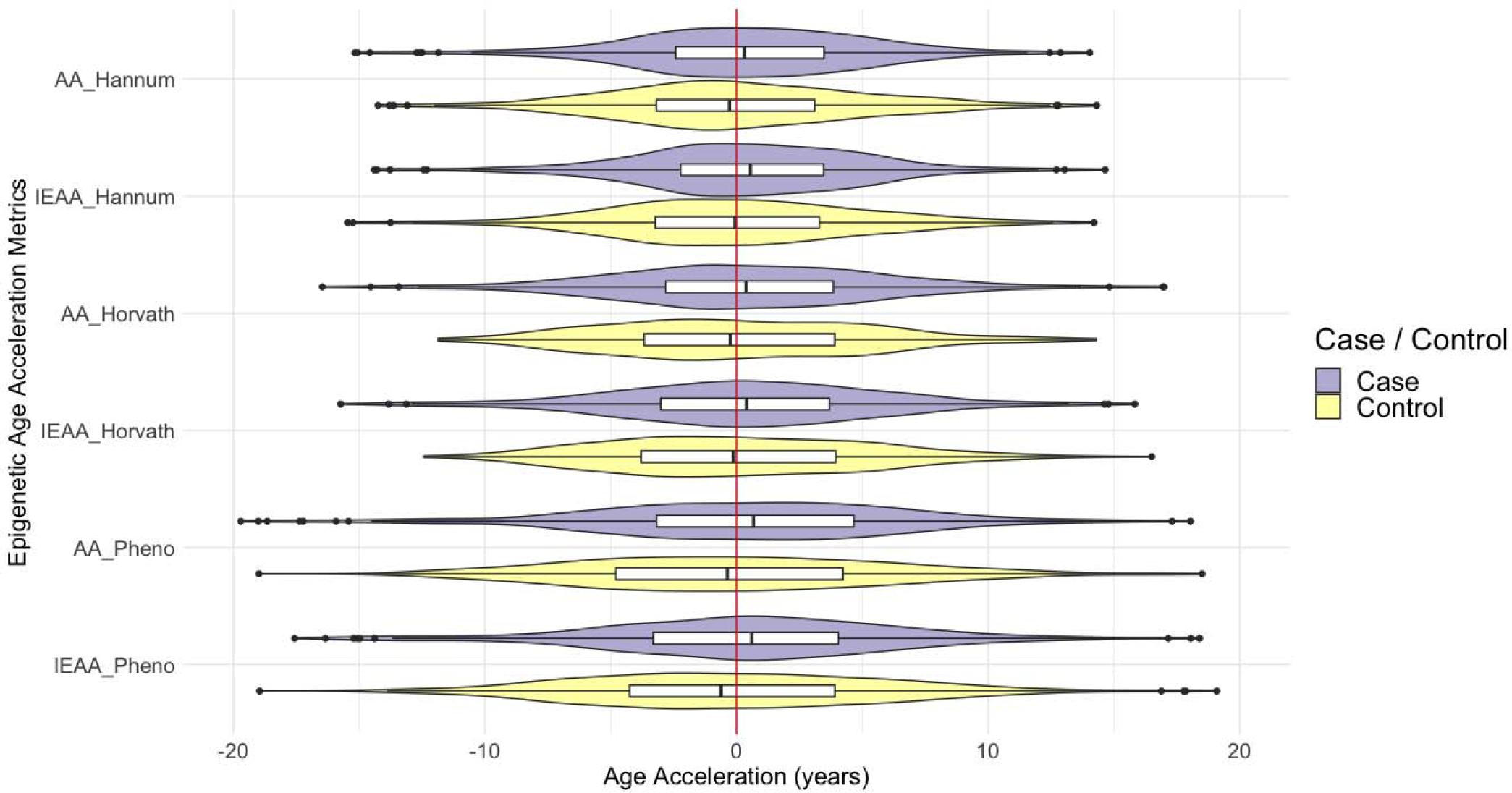
Distributions of 6 age acceleration metrics among pancreatic cancer cases and their matched controls. Legends: Violin plots of the distributions of three age acceleration (AA) and three intrinsic age acceleration (IEAA) metrics estimated by Hannum’s clock, Horvath’s clock, and PhenoAge among pancreatic cancer cases and their matched controls.

### DNAm age acceleration and pancreatic cancer risk

Conditional logistic regression was performed to examine the associations between epigenetic age acceleration and pancreatic cancer risk. All statistical models adjusted for matching factors, including cohorts (HPFS, NHS, or PHS), age, date of blood draw, race, and smoking status, in addition to BMI as a covariate. Participants were grouped into quartiles according to the distribution of each epigenetic AA and IEAA metric among controls. Quartiles 1 and 2 are comprised of pancreatic cancer cases and their matched controls with values of epigenetic AA or IEAA metrics less than zero (decelerated aging). Conversely, quartiles 3 and 4 are comprised of participants with values of epigenetic AA or IEAA metrics greater than zero (accelerated aging). Comparing participants in the highest quartile of age acceleration metrics (the cutoffs for these metrics varied) to those in the lowest quartile (the cutoffs for these metrics varied), the pancreatic cancer risks were significantly increased by 67% to 83% with minimal of 7- to 9-years accelerated ages for Hannum and PhenoAge AA or IEAA metrics. Comparing the highest to the lowest quartile, pancreatic cancer risks were increased by 28% for Horvath AA and IEAA metrics although these associations were not statistically significant (Figure 4); however, for those measures, the risks were higher when comparing participants in the third quartile to the lowest quartile (Horvath AA: odds ratio [OR] = 1.46; 95% 95% confidence interval [CI] = 0.95 to 2.24; Horvath IEAA: OR = 1.89; 95% CI = 1.23 to 2.96). Except for Hovarth AA and IEAA metrics, there were significant dose-response trends, such that higher age accelerations were associated with higher pancreatic cancer risks, but the relationship appeared nonlinear. (Figure 4)

**Figure 4:**
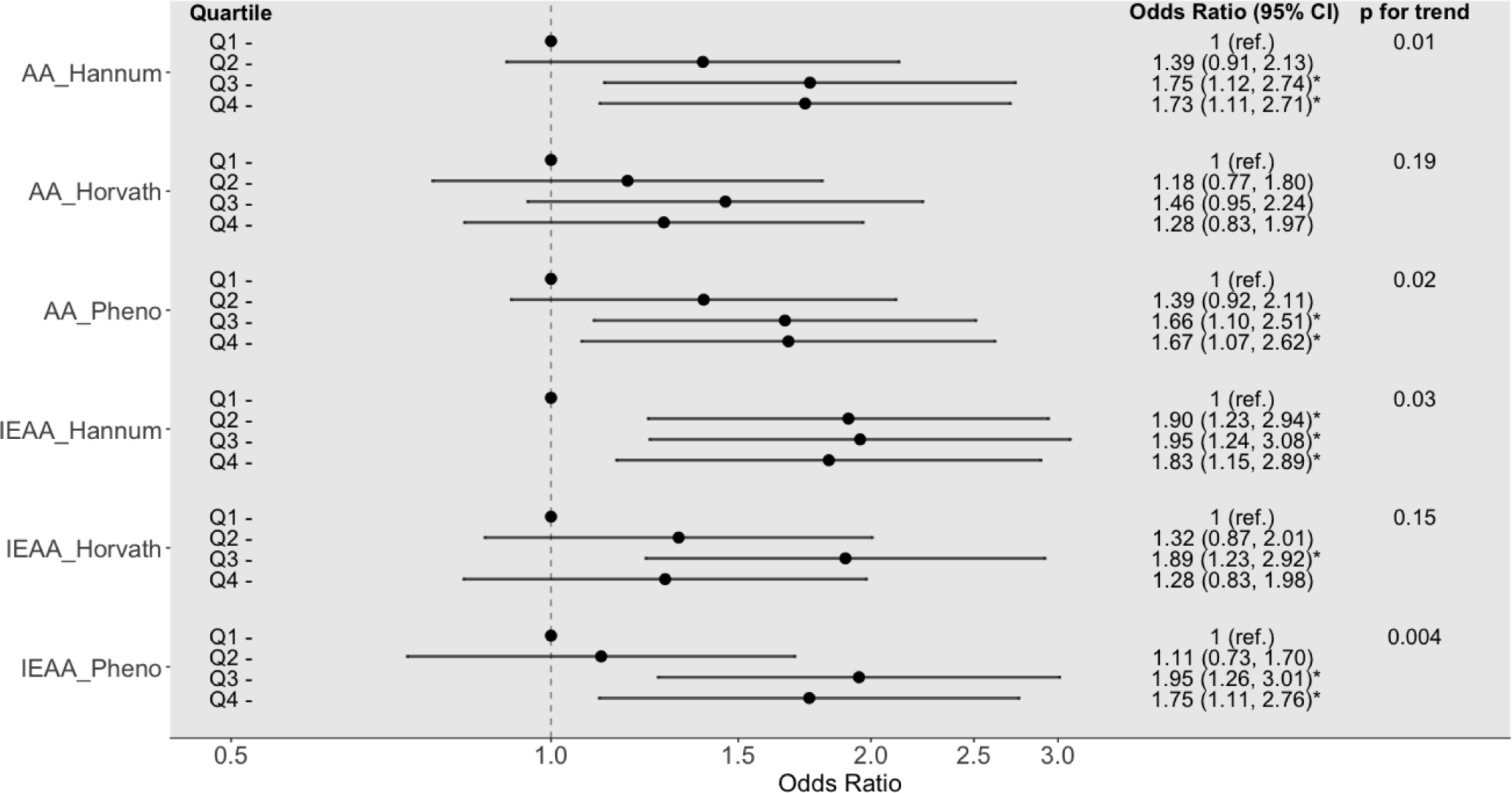
Forest plot of the associations between age acceleration metrics (per quintile) and pancreatic cancer risk. Legends: AA = epigenetic age acceleration metric; CI = confidence interval; IEAA = intrinsic epigenetic age acceleration metric; ref. = reference group. Please refer to Table footnotes for quartile cut-offs.

Stratified analyses were conducted to explore whether the associations between DNAm age acceleration and pancreatic cancer risk differed by participants’ characteristics. In these analyses, the associations were heterogeneous, varying by participants’ characteristics and by epigenetic AA or IEAA metrics (Table 2), although some general trends were observed. Specifically, the associations were larger in male than female for Hannum IEAA and Horvath AA or IEAA metrics, but the associations were similar comparing male to female for Hannum AA and PhenoAge AA or IEAA metrics. Compared to the elderly (age >65 years), the associations were generally smaller for Hannum and PhenoAge AA or IEAA in the younger age group (age ≤65 years). In contrast, the Horvath AA or IEAA metrics were more strongly associated with risk in the younger age group compared to the elderly. For all epigenetic AA and IEAA metrics, the associations were larger in overweight or obese participants (BMI ≥25 kg/m^2^) than in normal or underweight participants. These results are mostly consistent with the results when analyses were conducted using quartiles of epigenetic age acceleration metrics (Supplemental Tables 2-5), except for the stratified analyses by smoking status. Specifically, strong positive associations (with wide CIs) between all AA or IEAA metrics and pancreatic cancer risk were observed among current smokers comparing participants in the higher quartiles to the lowest quartile (ORs ranged from 1.49 to 3.55). There were fewer current smokers in the lowest quartile (larger decelerated epigenetic ages) than higher quartiles, especially for the PhenoAge AA or IEAA metrics (15% and 17% of smokers in quartile 1, respectively). Thus, the median of all three epigenetic AA and three IEAA metrics were above zero for both pancreatic cancer cases and matched controls.

**Table 2.** Risks of pancreatic cancer comparing individuals with epigenetically accelerated age to those with decelerated age, stratified by baseline characteristics

|  | **Hannum AA** | **Horvath AA** | **PhenoAge AA** | **Hannum IEAA** | **Horvath IEAA** | **PhenoAge IEAA** |
| --- | --- | --- | --- | --- | --- | --- |
|  | OR (95% CI)<sup>a</sup> | OR (95% CI)<sup>a</sup> | OR (95% CI)<sup>a</sup> | OR (95% CI)<sup>a</sup> | OR (95% CI)<sup>a</sup> | OR (95% CI)<sup>a</sup> |
| **Overall** | 1.33 (0.99, 1.80) | 1.31 (0.97, 1.78) | 1.30 (0.96, 1.74) | 1.32 (0.97, 1.81) | 1.44 (1.07, 1.93)* | 1.41 (1.03, 1.92)* |
| **Sex** |  |  |  |  |  |  |
| Female | 1.29 (0.85, 1.97) | 1.16 (0.76, 1.78) | 1.21 (0.80, 1.82) | 1.18 (0.74, 1.86) | 1.26 (0.83, 1.91) | 1.36 (0.87, 2.12) |
| Male | 1.36 (0.89, 2.09) | 1.47 (0.96, 2.25) | 1.39 (0.90, 2.14) | 1.46 (0.95, 2.25) | 1.64 (1.07, 2.52)* | 1.47 (0.95, 2.27) |
| **Age at blood draw** |  |  |  |  |  |  |
| ≤ 65 year | 1.25 (0.87, 1.80) | 1.52 (1.05, 2.21)* | 1.35 (0.95, 1.94) | 1.19 (0.82, 1.75) | 1.56 (1.08, 2.23)* | 1.48 (1.02, 2.14)* |
| > 65 year | 1.70 (0.92, 3.13) | 1.08 (0.59, 1.97) | 1.50 (0.80, 2.81) | 1.71 (0.92, 3.16) | 1.25 (0.68, 2.28) | 1.51 (0.76, 3.00) |
| **Smoking status** |  |  |  |  |  |  |
| Never | 1.40 (0.88, 2.23) | 1.96 (1.18, 3.25)* | 1.59 (0.99, 2.56) | 1.48 (0.90, 2.45) | 2.08 (1.25, 3.44)* | 2.08 (1.22, 3.55)* |
| Former | 1.17 (0.69, 1.97) | 0.82 (0.50, 1.34) | 0.97 (0.59, 1.57) | 1.31 (0.78, 2.21) | 0.86 (0.53, 1.41) | 1.09 (0.66, 1.79) |
| Current | 1.07 (0.48, 2.40) | 0.98 (0.43, 2.27) | 0.83 (0.33, 2.07) | 1.01 (0.43, 2.36) | 1.76 (0.71, 4.37) | 1.19 (0.51, 2.78) |
| **BMI** |  |  |  |  |  |  |
| < 25 kg/m<sup>2</sup> | 1.46 (0.72, 2.96) | 1.29 (0.66, 2.55) | 1.21 (0.62, 2.36) | 0.94 (0.49, 1.83) | 1.34 (0.70, 2.57) | 1.14 (0.59, 2.19) |
| ≥ 25 kg/m<sup>2</sup> | 1.51 (0.85, 2.66) | 1.31 (0.71, 2.40) | 1.63 (0.87, 3.04) | 1.47 (0.83, 2.61) | 1.76 (0.94, 3.31) | 1.57 (0.83, 3.00) |

AA = epigenetic age acceleration metric; CI = confidence interval; IEAA = intrinsic epigenetic age acceleration metric; OR = odds ratio.
<sup>a</sup> All conditional logistic regression models were adjusted for BMI, except for the analysis by BMI categories.
\* p-value ≤ 0.05

### DNAm age acceleration and time to diagnosis

To examine whether the epigenetic clocks were affected by clinically occult cancer, we conducted stratified analyses by time to cancer diagnosis (≤10, 10-15, or >15 years). After accounting for the matching factors and BMI, epigenetic AA and IEAA metrics showed different trends in associations with cancer risk across categories of time to cancer diagnosis (Table 3). As time to diagnosis increased, the ORs of pancreatic cancer for the Hannum AA and Horvath AA or IEAA metrics trended upwards, while the ORs for the PhenoAge AA or IEAA and Hannum IEAA metrics trended downward. Additionally, stratified analyses by time to cancer diagnosis revealed that Hannum IEAA was significantly positively associated with cancer risk among participants who were within 10 years or less to cancer diagnosis (Hannum IEAA Q3 vs. Q1 OR□=□2.83, 95% CI = 1.08 to 7.39, *P*□= 0.033). All age acceleration metrics, except for Horvath AA, showed significant positive associations with cancer risk among those who were 15 years or more to cancer diagnosis (Table 3)

**Table 3.** Epigenetic age acceleration metrics (per quintile) and pancreatic cancer risk stratified by time to cancer diagnosis

| Epigenetic age acceleration metrics | Quartile comparison | ToDx ≤ 10 years<br>(case/control *n*: 133/141) |  | ToDx 10-15 years<br>(case/control *n*: 91/96) |  | ToDx >15 years<br>(case/control *n*: 157/158) |  |
| --- | --- | --- | --- | --- | --- | --- | --- |
|  |  | OR (95% CI)<sup>a</sup> | *P* | OR (95% CI)<sup>a</sup> | *P* | OR (95% CI)<sup>a</sup> | *P* |
| AA_Hannum<sup>b</sup> | Q2 vs. Q1 | 1.26 (0.57, 2.79) | 0.57 | 1.01 (0.44, 2.34) | 0.98 | 1.69 (0.86, 3.34) | 0.13 |
|  | Q3 vs. Q1 | 2.31 (0.95, 5.64) | 0.07 | 1.10 (0.43, 2.80) | 0.84 | 1.70 (0.88, 3.28) | 0.11 |
|  | Q4 vs. Q1 | 1.27 (0.54, 2.97) | 0.58 | 1.74 (0.72, 4.20) | 0.22 | 2.25 (1.10, 4.59) | 0.03 |
|  | *P* for trend |  | 0.65 |  | 0.18 |  | 0.03 |
| AA_Horvath<sup>b</sup> | Q2 vs. Q1 | 1.33 (0.63, 2.83) | 0.45 | 1.07 (0.47, 2.44) | 0.88 | 1.20 (0.59, 2.44) | 0.62 |
|  | Q3 vs. Q1 | 1.28 (0.59, 2.77) | 0.53 | 1.72 (0.76, 3.89) | 0.19 | 1.57 (0.76, 3.27) | 0.23 |
|  | Q4 vs. Q1 | 0.75 (0.36, 1.56) | 0.44 | 1.38 (0.60, 3.18) | 0.45 | 2.11 (0.96, 4.63) | 0.06 |
|  | *P* for trend |  | 0.33 |  | 0.3 |  | 0.04 |
| AA_PhenoAge<sup>b</sup> | Q2 vs. Q1 | 1.83 (0.86, 3.91) | 0.12 | 1.37 (0.59, 3.19) | 0.46 | 1.12 (0.58, 2.17) | 0.74 |
|  | Q3 vs. Q1 | 1.56 (0.78, 3.15) | 0.21 | 1.10 (0.49, 2.48) | 0.81 | 2.08 (1.01, 4.31) | 0.048 |
|  | Q4 vs. Q1 | 1.71 (0.74, 3.97) | 0.21 | 2.10 (0.87, 5.05) | 0.10 | 1.46 (0.70, 3.05) | 0.31 |
|  | *P* for trend |  | 0.29 |  | 0.16 |  | 0.15 |
| IEAA_Hannum<sup>b</sup> | Q2 vs. Q1 | 2.43 (0.93, 6.32) | 0.07 | 1.39 (0.63, 3.07) | 0.41 | 2.07 (1.07, 4.04) | 0.03 |
|  | Q3 vs. Q1 | 2.83 (1.08, 7.39) | 0.03 | 1.44 (0.56, 3.72) | 0.45 | 1.81 (0.90, 3.68) | 0.10 |
|  | Q4 vs. Q1 | 1.69 (0.63, 4.57) | 0.3 | 1.71 (0.73, 4.02) | 0.22 | 2.08 (1.01, 4.29) | 0.05 |
|  | *P* for trend |  | 0.78 |  | 0.24 |  | 0.07 |
| IEAA_Horvath<sup>b</sup> | Q2 vs. Q1 | 1.70 (0.79, 3.68) | 0.18 | 0.60 (0.24, 1.48) | 0.27 | 1.81 (0.91, 3.61) | 0.09 |
|  | Q3 vs. Q1 | 2.20 (0.96, 5.04) | 0.06 | 1.59 (0.69, 3.67) | 0.27 | 2.19 (1.07, 4.47) | 0.03 |
|  | Q4 vs. Q1 | 0.74 (0.35, 1.59) | 0.44 | 1.22 (0.53, 2.80) | 0.63 | 2.53 (1.12, 5.72) | 0.03 |
|  | *P* for trend |  | 0.28 |  | 0.3 |  | 0.02 |
| IEAA_PhenoAge<sup>b</sup> | Q2 vs. Q1 | 2.00 (0.93, 4.26) | 0.07 | 0.53 (0.23, 1.22) | 0.13 | 1.20 (0.60, 2.41) | 0.61 |
|  | Q3 vs. Q1 | 1.78 (0.84, 3.78) | 0.13 | 1.54 (0.67, 3.54) | 0.31 | 2.94 (1.32, 6.54) | 0.008 |
|  | Q4 vs. Q1 | 2.17 (0.96, 4.91) | 0.06 | 1.14 (0.49, 2.66) | 0.75 | 1.70 (0.76, 3.79) | 0.20 |
|  | *P* for trend |  | 0.10 |  | 0.37 |  | 0.13 |

AA = epigenetic age acceleration metric; IEAA = intrinsic epigenetic age acceleration metric; *n* = sample size; OR = odds ratio; Q1 = quartile 1; Q2 = quartile 2; Q3 = quartile 3; Q4 = quartile 4; ToDx = time to cancer diagnosis.
<sup>a</sup> All conditional logistic regression models were adjusted for BMI.
<sup>b</sup> Quartile cut-offs: Q1: <-3.2 for AA\_Hannum, < -3.7 for AA\_Horvath, < -4.8 for AA\_Pheno, < -3.2 for IEAA\_Hannum, for <-3.8 IEAA\_Horvath and <-4.2 for IEAA\_Pheno; Q2: -3.2 to -0.31 for AA\_Hannum, -3.7 to -0.21 for AA\_Horvath, -4.8 to -0.41for AA\_Pheno, -3.2 to -0.11 for IEAA\_Hannum, -3.8 to -0.11 for IEAA\_Horvath and -4.2 to -0.61 for IEAA\_Pheno; Q3: -0.3 to 3.1 for AA\_Hannum, -0.2 to 3.9 for AA\_Horvath, -0.4 to 4.2 for AA\_Pheno, -0.1 to 3.3 for IEAA\_Hannum, -0.1 to 3.9 for IEAA\_Horvath and -0.6 to 3.9 for IEAA\_Pheno; Q4: $\geq 3.1$ for AA\_Hannum, $\geq 3.9$ for AA\_Horvath, $\geq 4.2$ for AA\_Pheno, $\geq 3.3$ for IEAA\_Hannum, $\geq 3.9$ for IEAA\_Horvath and $\geq 3.9$ for IEAA\_Pheno.

### DNAm age acceleration and pancreatic cancer survival

We conducted case-only analyses to examine the associations between DNAm age acceleration and pancreatic cancer survival. Hazard ratios for pancreatic cancer survival were similar across all epigenetic AA and IEAA metrics. Overall, we observed little evidence of any significant association between pancreatic cancer survival and any of the epigenetic AA or IEAA metrics (Table 4).

**Table 4.** Associations between epigenetic age acceleration metrics (higher quintile vs. the lowest quintile) and pancreatic cancer survival in pancreatic cancer cases

| Epigenetic age acceleration metrics | Quartiles<sup>a</sup> | N | HR (95% CI)<sup>b</sup> | P-value | P for trend |
| --- | --- | --- | --- | --- | --- |
| AA_Hannum | Q1 | 69 | 1 (ref.) |  | 0.96 |
|  | Q2 | 83 | 1.36 (0.97, 1.92) | 0.08 |  |
|  | Q3 | 100 | 1.24 (0.89, 1.73) | 0.21 |  |
|  | Q4 | 82 | 1.07 (0.77, 1.50) | 0.68 |  |
| AA_Horvath | Q1 | 61 | 1 (ref.) |  | 0.71 |
|  | Q2 | 79 | 1.21 (0.87, 1.68) | 0.26 |  |
|  | Q3 | 95 | 1.17 (0.86, 1.61) | 0.32 |  |
|  | Q4 | 97 | 0.94 (0.67, 1.32) | 0.72 |  |
| AA_PhenoAge | Q1 | 63 | 1 (ref.) |  | 0.22 |
|  | Q2 | 84 | 1.21 (0.86, 1.69) | 0.27 |  |
|  | Q3 | 97 | 1.10 (0.80, 1.52) | 0.56 |  |
|  | Q4 | 91 | 1.30 (0.92, 1.82) | 0.13 |  |
| IEAA_Hannum | Q1 | 62 | 1 (ref.) |  | 0.96 |
|  | Q2 | 83 | 1.45 (1.02, 2.05) | 0.04 |  |
|  | Q3 | 115 | 1.29 (0.91, 1.83) | 0.16 |  |
|  | Q4 | 74 | 1.13 (0.79, 1.61) | 0.5 |  |
| IEAA_Horvath | Q1 | 53 | 1 (ref.) |  | 0.73 |
|  | Q2 | 95 | 1.19 (0.85, 1.67) | 0.3 |  |
|  | Q3 | 96 | 1.13 (0.82, 1.56) | 0.46 |  |
|  | Q4 | 88 | 1.09 (0.76, 1.56) | 0.64 |  |
| IEAA_PhenoAge | Q1 | 63 | 1 (ref.) |  | 0.65 |
|  | Q2 | 72 | 1.15 (0.81, 1.63) | 0.44 |  |
|  | Q3 | 110 | 1.16 (0.84, 1.59) | 0.37 |  |
|  | Q4 | 89 | 1.10 (0.78, 1.54) | 0.58 |  |

AA = epigenetic age acceleration metric; CI = confidence interval; IEAA = intrinsic epigenetic age acceleration metric; *n* = sample size; HR = hazard ratio; ref. = reference group.
<sup>a</sup>Quartile cut-offs: Q1: <-3.2 for AA\_Hannum, <-3.7 for AA\_Horvath, <-4.8 for AA\_Pheno, <-3.2 for IEAA\_Hannum, for <-3.8 IEAA\_Horvath and <-4.2 for IEAA\_Pheno; Q2: -3.2 to -0.31 for AA\_Hannum, -3.7 to -0.21 for AA\_Horvath, -4.8 to -0.41 for AA\_Pheno, -3.2 to -0.11 for IEAA\_Hannum, -3.8 to -0.11 for IEAA\_Horvath and -4.2 to -0.61 for IEAA\_Pheno; Q3: -0.3 to 3.1 for AA\_Hannum, -0.2 to 3.9 for AA\_Horvath, -0.4 to 4.2 for AA\_Pheno, -0.1 to 3.3 for IEAA\_Hannum, -0.1 to 3.9 for IEAA\_Horvath and -0.6 to 3.9 for IEAA\_Pheno; Q4: ≥ 3.1 for AA\_Hannum, ≥ 3.9 for AA\_Horvath, ≥ 4.2 for AA\_Pheno, ≥ 3.3 for IEAA\_Hannum, ≥ 3.9 for IEAA\_Horvath, and ≥ 3.9 for IEAA\_Pheno.
<sup>b</sup>Cox proportional hazard regression models were adjusted for cohorts, age at blood draw, race, date of blood draw, smoking status and BMI.

## Discussions

Our results indicate DNAm age acceleration is associated with an increased risk of pancreatic cancer that is independent of several key risk factors for cancer including chronological age. We found positive associations for all three epigenetic clocks (Hannum, Horvath, and PhenoAge), with stronger associations for intrinsic epigenetic age acceleration (IEAA) metrics suggesting that blood cell composition is a weak confounder in our analyses. Moreover, our findings suggest that the association between DNAm age acceleration and pancreatic cancer risk is possibly in a nonlinear, dose-response manner.

While prior studies have examined the epigenetic clock associations with cancer risk (12-16), only one study examined PhenoAge clock in relation to breast cancer risk (16) and another assessed nonlinear relationships between Hannum or Horvath IEAA and all cancer incidence and mortality (13). Analyzing multiple epigenetic age acceleration over time, the latter study found complex linear and nonlinear, dynamic time-dependent relationship with all cancer incidence and mortality (13). We did not find a significant association between age acceleration and pancreatic cancer survival. It should be noted that the prognosis of pancreatic cancer is poor (> 90% of pancreatic cancer cases died within 2.5 years after diagnosis) so associations between age acceleration and pancreatic cancer survival would be hard to detect, if they exist (Supplemental Figure 1)

The three epigenetic clocks have only a few CpGs in common, but the AA or IEAA metrics were highly correlated with each other in our study sample. The correlations between these epigenetic AA or IEAA metrics were moderate to high (*r* ranged from 0.39 to 0.98) in a case-cohort study of breast cancer risk (16). Unlike previous studies (none of these studies included pancreatic cancer cases) (12, 15, 16), the correlations between the epigenetic age estimated by the three epigenetic clocks and chronological age were only moderate in our study sample. The PhenoAge clock showed the lowest correlation to chronological age, which is not surprising because PhenoAge clock was trained on age-related and disease phenotypes in addition to chronological age (24). While different cancers are biologically distinct from each other, epigenetic age acceleration may not be cancer specific. The process of carcinogenesis is almost universally associated with both inflammation and activation of immune senescence pathways (25, 26). The fact that our findings also are consistent with those of the only prior study that examined the associations between age acceleration estimated by Hannum, Horvath, or PhenoAge clocks and risk of breast cancer (16) may also suggest that age acceleration reflects a more general cancer-associated biological process.

We observed heterogeneous associations, varying by participants’ characteristics, time to diagnosis, and AA or IEAA metrics in the stratified analyses. These findings suggest that the three epigenetic clocks (Hannum, Horvath, and PhenoAge) capture different aspects of biological aging and abnormal changes to cancer development, although the biological implications of the epigenetic clock are not well understood. Nonetheless, studies have consistently shown that BMI or obesity is positively associated with Hannum and Horvath age acceleration metrics (27-30), which support our findings with regard to the associations with pancreatic cancer that were larger in overweight or obese participants than in normal or underweight participants. Since studies have identified many common genetic variants associated with BMI or obesity (31), these findings point to a genetic component of these epigenetic clocks (12). Additionally, our stratified analyses showed that associations between all AA or IEAA metrics and pancreatic risk are modified by smoking status, suggesting that these epigenetic clocks are affected by lifestyle factors including smoking (27, 30, 32). Compared to 5 different DNAm aging clocks (including Hannum and Horvath), PhenoAge clock was shown to have stronger associations with all-cause mortality, smoking status, leukocyte telomere length, naïve CD8+ T cells and CD4+ T cells (24).

Lastly, our results overall suggest that Hannum and PhenoAge IEAA metrics may be more suitable for evaluating the associations with pancreatic cancer risks than Horvath clock, because smaller associations and opposite trends in associations with cancer risk across categories of time to cancer diagnosis were shown for Horvath IEAA metrics compared to Hannum and PhenoAge IEAA in our study sample. Age-related DNAm signatures may be influenced by tissue’s cell composition, which is altered with age and may partially mask age acceleration associations to disease risk. IEAA metrics are not confounded by the blood cell compositions by definition.

Our study has several strengths. We employed a prospective study design by using prediagnostic bloods of incidence pancreatic cancer cases and matched controls from three large cohort studies in the United States. Our analyses minimize confounding through matching and statistical adjustments. Although we find evidence that age acceleration is associated with increased risk of pancreatic cancer, our study has several limitations. First, our findings do not provide mechanistic insights into how these age-related changes influence pancreatic cancer risks. Second, our study population is comprised of mostly Caucasians so our results may not be generalizable to other races or ethnicity. Lastly, our stratified analyses may have spurious findings due to multiple testing and small sample sizes in some subgroups.

In sum, our study is first prospective study to examine Hannum, Horvath, and PhenoAge epigenetic clocks in relation to pancreatic cancer risk or survival, and we find that DNAm age acceleration is associated with an increased risk of pancreatic cancer in a nonlinear, dose-response manner. Epigenetic IEAA metrics may be used to improve pancreatic cancer risk prediction when used in concert with known disease risk factors. Little is known regarding the trajectories of these epigenetic clock in relation to cancer risk. To build evidence base to support epigenetic age as a biomarker for cancer early detection, future prospective studies should evaluate the changes in DNAm age (ideally at multiple time points) relating to cancer development.

## Data Availability

All data from this study have been deposited in dbGAP and will be available on January 3, 2020 [DNA Methylation Markers and Pancreatic Cancer Risk in 3 Cohort Studies (NHS, PHS, HPFS) phs001917.v1.p1].

https://www.ncbi.nlm.nih.gov/projects/gapprev/gap/cgi-bin/study.cgi?study_id=phs001917.v1.p1

